# Investigating the potential benefit that requiring travellers to self-isolate on arrival may have upon the reducing of case importations during international outbreaks of influenza, SARS, Ebola virus disease and COVID-19

**DOI:** 10.1101/2020.10.02.20205757

**Authors:** Declan Bays, Emma Bennett, Thomas Finnie

## Abstract

With the advent of rapid international travel, disease can now spread between nations faster than ever. As such, when outbreaks occur in foreign states, pressure mounts to reduce the risk of importing cases to the home nation. In a previous paper, we developed a model to investigate the potential effectiveness of deploying screening at airports during outbreaks of influenza, SARS, and Ebola. We also applied the model to the current COVID-19 outbreak. This model simulated the testing of travellers (assumed not to be displaying symptoms prior to boarding their flight) as they arrived at their destination. The model showed that the reduction in risk of case importation that screening alone could deliver was minimal across most scenarios considered, with outputs indicating that screening alone could detect at most 46.4%, 12.9%, and 4.0% of travellers infected with influenza, SARS and Ebola respectively, while the model also reported a detection rate of 12.0% for COVID-19. In this paper, we present a brief modification to this model allowing us to assess the added impact that quarantining incoming travelers for various periods may have on reducing the risk of case importation. Primary results show that requiring all travellers to undergo 5 days of self-isolation on arrival, after which they are tested again, has the potential to increase rates of detection to 100%, 87.6%, 81.7% and 41.3% for travellers infected with influenza, SARS, COVID-19 and Ebola respectively. Extending the period of self-isolation to 14 days increases these potential detection rates to 100%, 100%, 99.5% and 91.8% respectively.

## Introduction

In a previous paper[1], we described an adaptable model which could be used to assess the effectiveness of implementing a strict screening policy at airports during international disease outbreaks. We subsequently used this model to evaluate the probability that screening would be able to detect infected travellers on arrival across a range of scenarios, given that they were well enough to fly (i.e. not symptomatic when they boarded their flight). These scenarios varied according to the type of disease travellers were infected with, the length of window within which travellers could have become infected, and whether travellers took a short, medium or long-haul flight.

The outcome of this work was that in the best-case scenario screening has the capability to detect 46.4%, 12.9%, 12.0% and 4.0% of travellers that have been infected with influenza, SARS, COVID-19 and Ebola respectively. These values, being as low as they are, seem to indicate that the impact of screening alone may not deliver sufficient effect to reduce the risk of importation to levels deemed sufficient by local authorities. The main reason for the reported success rates being so low is that if one considers only the infected travellers that are not symptomatic prior to flying (i.e. not detectable to testing), then their flight only provides a small additional window for travellers to incubate. It is only those that become symptomatic in this short window that entry screening would detect. Following this argument, one may logically suggest that rates of detection could be improved if this window were extended by means comparable to requiring all incoming travellers to undergo observation for some given period post-arrival (in the form of self-isolation or other similar means). After this, travellers could then be given an additional test, whereby only if their results come back negative would they be released into the wider population.

In this paper, we investigate whether the addition of requiring arriving travellers to undergo such a self-isolation period to the border screening process might improve detection rates enough to present an attractive alternative during such outbreaks. We achieve this by way of modifying the model presented in the original paper[1] to allow for this additional period of isolation. As such, this paper should be seen as an addendum to the previous work. Results reported are again obtained through the use of Monte Carlo simulation, under the same set of assumptions as before.

## Assumptions

This work is based heavily upon [1]. As such, the basic assumptions remain unchanged. We present these here:

- The time each traveller became infected before their flight, −t_0_ is sampled from a uniform distribution, *D*_exp_, over the intervals [0, 72], [0, 168] or [0, 336] depending on the outbreak scenario being considered.
- Travellers that have become symptomatic prior to boarding their flight do not fly.
- All persons travelling only take directs flight from country A to their destination, country B.
- The flight time for each traveller is sampled from a uniform distribution, *D*_flight_, over the intervals [3,5], [7,9] or [11,13] depending on whether a short, medium or long-haul flight is being considered.
- All travellers that fly to country B are tested on arrival.
- All travellers that enter quarantine are tested at the end of their quarantine period.
- Testing only detects travellers that are symptomatic; prior to symptom onset they are not detectable.
- Screening does not produce false negative.
- The number of infected persons remains constant throughout the simulation (transmission and death are neglected).
- Each disease is characterised solely by its incubation period distribution, *D*_inc_.
- The incubation period distributions of each of the considered diseases are known or well approximated.
- Testing is 100% effective and detects all infected persons who have become symptomatic.
- Infected people do not attempt to “game” the system by concealing their symptomology.

## Methods

In this section, we recap the underlying structure and mechanics of the original model. We then explain how we have expanded the model to incorporate the additional self-isolation period under study. For a complete breakdown of the original model, we redirect the reader to the supplementary material included with the original work[1].

For each of the scenarios considered (described below), our model uses Monte Carlo methods to simulate a series of individuals, each being infected at a sampled time, −t_0_, before attempting to board a flight from country A to country B at time t=0. Each traveller is assigned an incubation time, T_inc_, after which they will be detectable by testing, and a flight time from country A to country B. These times are sampled from the distributions *D*_exp_, *D*_inc_ and *D*_flight_ respectively, each of which varies depending on the scenario being considered. The incubation distributions for influenza, SARS, COVID-19 and Ebola have been taken from the works [2]–[5] respectively, just as in [1]. To extend our previous work, travellers that successfully arrive into country B undetected must then undergo self-isolation for a fixed period, T_iso_. For each simulated individual, the model proceeds accordingly:

- If the incubation time is less than the time from infection to that person’s flight (T_inc_ < t_0_), they have become symptomatic before boarding their flight. Hence, they do not travel (either by being too sick or being picked up at exit screening); they exit the model being recorded as a *non-flier*.
- Or else, if their incubation time is less than the length of time from infection until that person’s flight lands in country B (T_inc_ < t_0_ + T_flight_), they have become symptomatic in transit. They will therefore be detected by testing administered upon arrival into country B; they are recorded as a *border detection*.
- Or else, the traveller has arrived into country B undetected. They then enter into self-isolation for a fixed period, T_iso_. If the incubation period is less than the time from infection until the end of the self-isolation period (T_inc_ < t_0_ + T_flight_ + T_iso_), then the traveller has become symptomatic during self-isolation. They will then be detected by the test administered on exit from self-isolation; they are recorded as a *self-isolation detection*.
- Else, their incubation time has exceeded the time taken for them to become infected, fly from country A to country B, undergo the self-isolation process and cross into country B (T_inc_ > t_0_ + T_flight_ + T_iso_); they then exit the model being recorded as a *non-detected case*.

We visualise this below:

**Figure 1:**
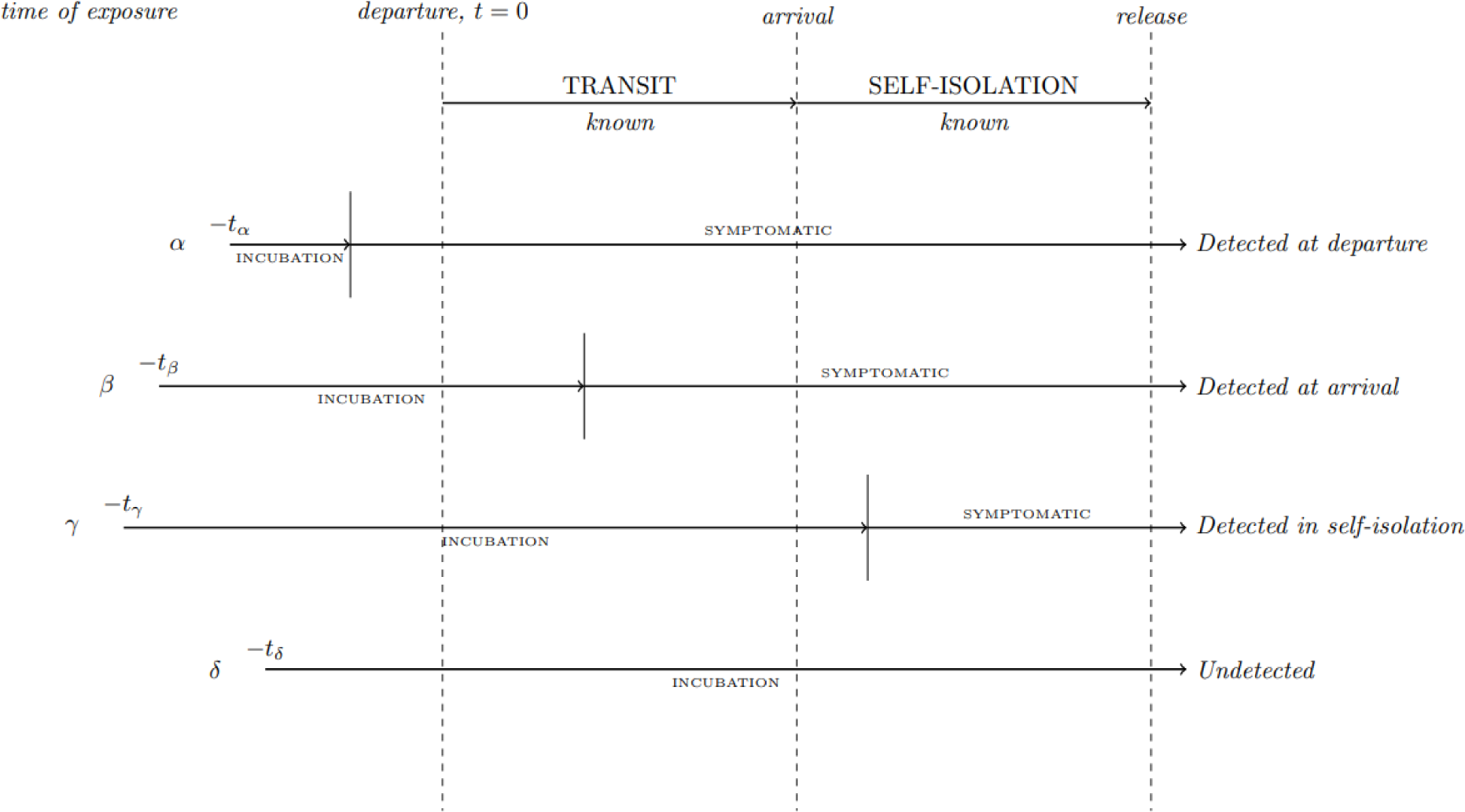
Depiction of how individuals in the model are evaluated. Traveller alpha becomes symptomatic prior to flight; recorded as a non-flier. Traveller beta becomes symptomatic in transit; recorded as an arrival detection. Traveller gamma become symptomatic in self-isolation; recorded as such. Traveller delta does not become symptomatic by the time self-isolation concludes; recorded as an undetected case.

This is then repeated across a range of outbreak scenarios, each of which considers 100,000 simulated travellers attempting to enter country B undetected. Scenarios are defined by a combination of disease, exposure range, flight time range and quarantine period. In the following we consider the diseases influenza, SARS, Ebola and COVID-19; each of these is captured using their respective incubation period distribution. Exposure ranges used are [0,72], [0,168] and [0,336] hours before flight. These then emulate scenarios where the traveller has been in country A (and hence at risk of infection) for a short period (e.g. due to a business trip), a medium period (e.g. on vacation), or more longer term (such as being a resident). Flight times are sampled from the ranges [3,5], [7,9] and [11,13] to model where travellers are required to take a short, medium or long-haul flight from country A to country B. Lastly, we consider quarantine periods which involves either 3, 5, 7, 10 or 14 days of self-isolation.

A Python package implementing the above model (which has been used to calculate the presented values in the next section) has been produced by the author and made openly available online[6].

## Results

The below plots display the outputs from our model. The raw outputted data has been included in the supplementary text file.

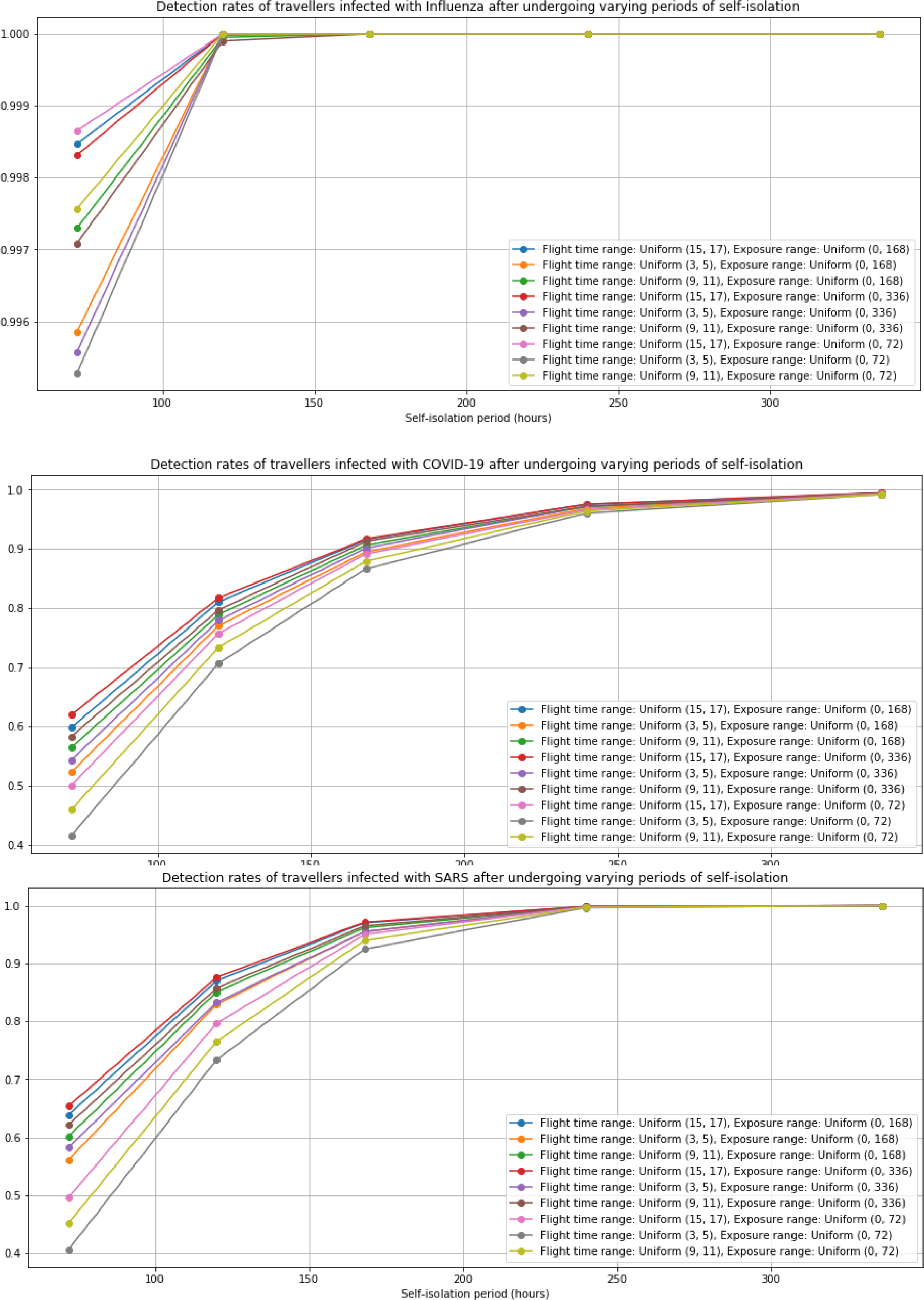
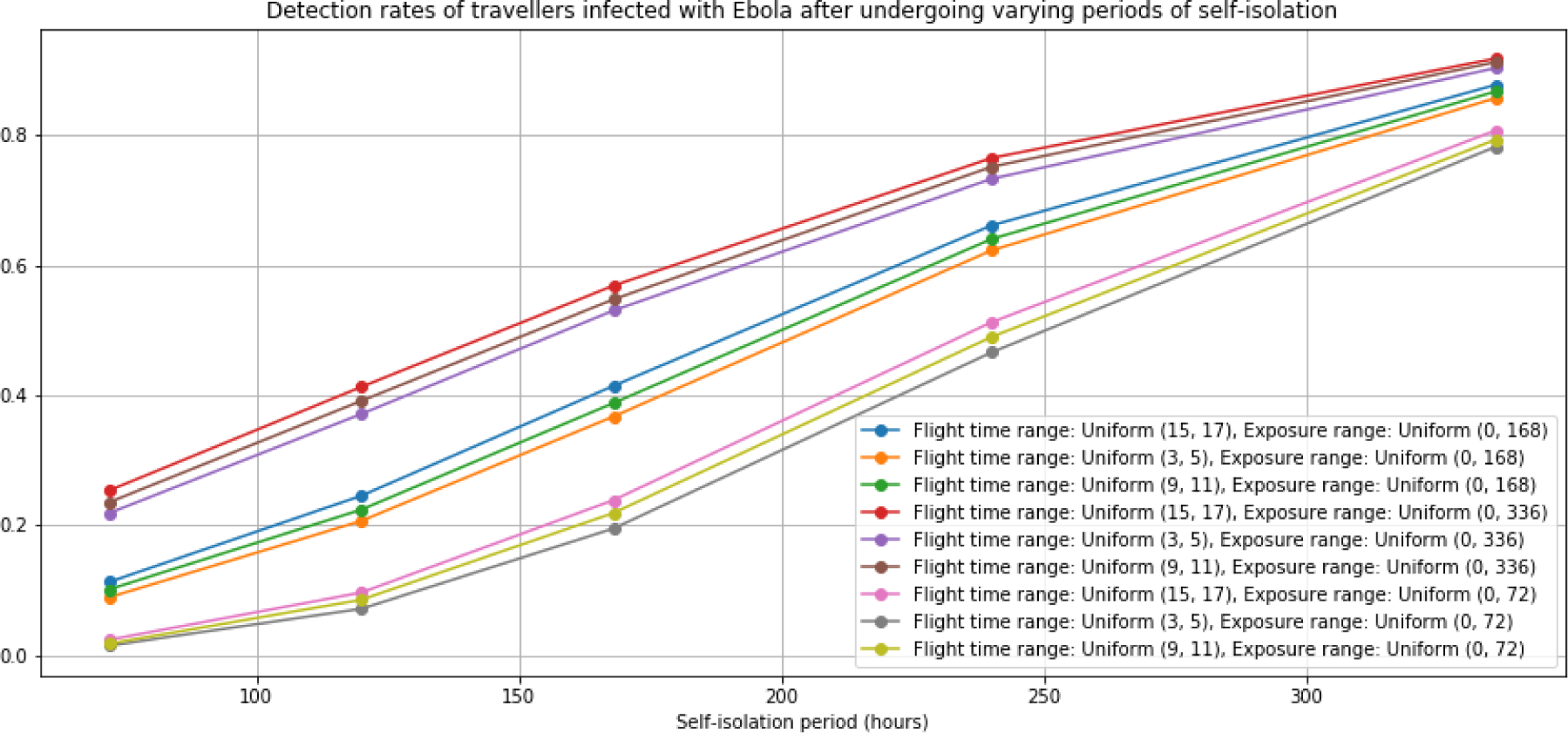

Recalling from our previous paper that detections rates, in lieu of any self-isolation period, was at most 46.4%, 12.9%, 4.0% and 12.0% for travellers infected with influenza, SARS, Ebola and COVID-19 respectively, the impact of imposing a self-isolation period can be clearly seen. Enforcing any of the considered periods of self-isolation acts to at least double the previously reported detection rate.

The most dramatic of our results, is that reported for influenza. The requirement for travellers to undertake as little as 3 days self-isolation acts to increase detection rates to over 99% across the board. Anything more than this pushes detection rates asymptotically closer to 100%.

The similar shapes of detection rate curves for SARS and COVID-19 are due to the similarity of their incubation period distributions. Note though how detection rates for SARS tops 90% across all scenarios once a 7-day self-isolation period is enforced, while the same self-isolation period for COVID-19 gives detection rates that congregate just shy of that. However, detection rates for both diseases are over 95% for all scenarios once travellers undergo 10-days of self-isolation or longer. Again, the benefits of the imposed self-isolation period are obvious.

The effect of self-isolation on detection rates is much more drawn out for Ebola however. This is likely due to the long incubation periods that are associated with the disease. However, enforcing a 10-day self-isolation period does increase detection rates by at least 1100%, with outputs being reported in the range of 46.5 - 76.5% (depending on scenario), while a 14-day period concentrates these results into the more impressive range of 78.2 - 91.8%.

## Discussion

From what has been shown above, when compared to the results described in our previous work[1], it is clear that the addition of any of the considered self-isolation periods dramatically increases the rates of detection of infected travellers after arrival. This is likely because requiring travellers to self-isolate for some extended period allows them considerably more time to incubate than afforded by the flight alone, and thus makes them much more likely to become detectable by testing.

The time required to maximise detection rates varies by disease, due to differing incubation distributions. For instance, 3 days self-isolation is sufficient to increase the detection rate of travellers infected with influenza from 46.4% (in the best-case scenario) to over 99.5% in all considered scenarios. While for Ebola, a disease which has a much longer average incubation period, the considered self-isolation periods have a much slower effect. A 14-day self-isolation period results in detection rates of at least 78.2%; this is up from the 4.0% detection rate obtained in the best-case scenario when no self-isolation was being enforced. The results for SARS and COVID-19 fall somewhere in between these two extremes, having incubation distributions which come somewhere in the middle.

Our results also appear to show that the amount of time travellers spend in transit (i.e. short, medium or long-haul flight) seems to impact less on detection rates as the length of self-isolation increases. This is not so true for Ebola, although initially it does have a “noticeable” effect. This would be because as the amount of time spent in self-isolation increases, the difference between the amount of time spent travelling becomes negligible. The reason for Ebola’s exception, where this variation starts off making little difference, becomes more noticeable before becoming negligible again, is most likely due to the very small number of travellers that are initially being detected. Ebola’s incubation period distribution is very protracted, and therefore, the amount of time that the various flight types allow travellers to incubate for makes little difference in the shorter quarantine periods. However, as longer self-isolation periods are enforced, and larger proportions of travellers are detected, the additional amount of time the longer flight types allow for travellers to incubate starts to have a more pronounced effect. This continues until the effect of transit time becomes dwarfed by the effect that spending extended periods in isolation has on likelihood that infected travellers will incubate, and hence increases detection rates.

Lastly, one may also notice that points appear to start clustering in triples before converging as self-isolation periods increase (most clearly in plots for influenza and Ebola). The clustering appears to be in accordance with exposure ranges of each of the given scenarios. This is intuitive since travellers in each of the exposure scenarios will be infected on average at around the same of time. This then means that on average, they will incubate for similar amounts of time prior to boarding their flight. The flight times will of course vary across scenarios, but then accounts for the minor variation between the triples. As the amount of time spent in self-isolation increases, it will start to outweigh the effect of the different exposure scenarios, resulting in the convergence seen in the later results.

## Conclusion

In this paper we have described a minor adjustment to the Monte Carlo based model we described in previous work[1], in order to study the effect that requiring travellers to self-isolate may have on disease detection rates. While other authors have evaluated the possible impacts that screening travellers solely upon arrival may have on reducing the risk of importing various diseases[7]–[9], less research has been conducted into the effects of enforcing a period of self-isolation. Of the work that has been done to-date, the majority focuses only on the effects of adopting a quarantine policy for the current COVID-19 outbreak[10], [11]. In this paper, we aim not only to build upon our previous work, but to also extend it as a general tool for other diseases and outbreak contexts. Our model code is an extension of the package described previously[6]. This tool can therefore be used to assess the effectiveness of implementing border screening and self-isolation as a contingency method across a broad spectrum of scenarios. Using our model may also remove the need to reproduce significant amounts of work for marginally different disease scenarios, while providing public health teams with a flexible tool to assess the impact of implementing quarantine, and which can quickly be adjusted and run with minimal effort.

## Data Availability

Supplementary material is included at the end of the paper

