## Supplementary text 1 for "Investigating the potential benefit that requiring travellers to self-isolate on arrival may have upon the reducing of case importations during international outbreaks of influenza, SARS, Ebola virus disease and COVID-19"

*Investigating into the potential benefit of enforcing the quarantining of travellers at borders to reduce case importation during international outbreaks of influenza, SARS, Ebola virus disease and COVID-19 – Supplementary text*

*Model description*

Below, we present a thorough breakdown of the algorithm implemented by our model to determine the success rates of each of the proposed scenarios.

On initialisation, each simulated traveller is assigned an incubation period,$T_{\mathrm{inc}}$, a time of infection, $t_{0}$, and a flight time, $T_{\mathrm{flight}}$. Each of these are sampled from the distributions $D_{\text{exp}},D_{\text{inc}}$ and $D_{\text{flight}}$ respectively, which are determined by the currently considered scenario. Also given by the scenario is the self-isolation period, $T_{\mathrm{iso}}$, which is fixed. The traveller then progresses through the model according to the below, as they attempt to board their flight to country B:

- If $T_{\mathrm{inc}}<t_{0}$, we assume the infected person has become symptomatic prior to boarding their flight and are thus detectable. We then assume that, either by means of exit screening or the traveller being too ill to fly, this traveller does not make it onto their flight. These are then recorded as “*non-fliers*”.
- If $T_{\mathrm{inc}}>t_{0}$, the traveller proceeds to board their flight that will take $T_{\text{flight}}$ hours, where for each traveller, depending on the current scenario, this has sampled from a uniform distribution on the ranges [3,5], [9, 11] or [15, 17].
- If $T_{\mathrm{inc}}<t_{0} + T_{\mathrm{flight}}$, we assume the infected person has become symptomatic during their flight. They will thus be detectable by the testing that is administered to all incoming travellers on arrival; in such case, infected travellers are then removed from the model, being recorded as “*detected, arrival*”.
- If $T_{\mathrm{inc}}>t_{0} + T_{\mathrm{flight}}$, then the traveller enters self-isolation for a period of $T_{\text{iso}}$ hours, after which the traveller must undergo a second test
- If $T_{\mathrm{inc}} <t_{0} + T_{\mathrm{flight}} + T_{\mathrm{iso}}$, then the traveller is deemed to have become detectable while in self-isolation. The traveller will then be detected by this second test, being recorded as a “*detected, iso.*”.
- Else, if $T_{\mathrm{inc}} >t_{0} + T_{\mathrm{flight}} + T_{\mathrm{iso}}$, then the traveller remains undetected after the administering of the final test. This infected traveller has therefore made it through the entire process without being detected. They are therefore also recorded as an “*undetected*”

This process is then repeated 100,000 for each scenario, recording the success rate (found by taking the ratio of infected travellers that have been detected during the double testing process, over the total number of infected travellers that successfully travelled to Bermuda) for each combination of self-isolation period and flight range. Note that this success rate is also an approximation to the probability that infected travellers would be detected by the implemented screening process given that they successfully boarded their flight to country B.

*Results*

Here we present the raw results as outputted by our model for each of the considered scenarios:

| Disease name | Flight time range | Exposure time range | Self-isolation period (hours) | Detection rate |
| --- | --- | --- | --- | --- |
| Ebola | Uniform (3, 5) | Uniform (0, 72) | 72 | 0.01450145 |
| Ebola | Uniform (9, 11) | Uniform (0, 72) | 72 | 0.017491574 |
| Ebola | Uniform (15, 17) | Uniform (0, 72) | 72 | 0.023512116 |
| Ebola | Uniform (3, 5) | Uniform (0, 168) | 72 | 0.088798742 |
| Ebola | Uniform (9, 11) | Uniform (0, 168) | 72 | 0.101050348 |
| Ebola | Uniform (15, 17) | Uniform (0, 168) | 72 | 0.112425195 |
| Ebola | Uniform (3, 5) | Uniform (0, 336) | 72 | 0.218388684 |
| Ebola | Uniform (9, 11) | Uniform (0, 336) | 72 | 0.23508465 |
| Ebola | Uniform (15, 17) | Uniform (0, 336) | 72 | 0.253287691 |
| Ebola | Uniform (3, 5) | Uniform (0, 72) | 120 | 0.071134979 |
| Ebola | Uniform (9, 11) | Uniform (0, 72) | 120 | 0.085065104 |
| Ebola | Uniform (15, 17) | Uniform (0, 72) | 120 | 0.096182504 |
| Ebola | Uniform (3, 5) | Uniform (0, 168) | 120 | 0.206414674 |
| Ebola | Uniform (9, 11) | Uniform (0, 168) | 120 | 0.22375573 |
| Ebola | Uniform (15, 17) | Uniform (0, 168) | 120 | 0.245080356 |
| Ebola | Uniform (3, 5) | Uniform (0, 336) | 120 | 0.370892886 |
| Ebola | Uniform (9, 11) | Uniform (0, 336) | 120 | 0.391014257 |
| Ebola | Uniform (15, 17) | Uniform (0, 336) | 120 | 0.412567869 |
| Ebola | Uniform (3, 5) | Uniform (0, 72) | 168 | 0.19469947 |
| Ebola | Uniform (9, 11) | Uniform (0, 72) | 168 | 0.218558413 |
| Ebola | Uniform (15, 17) | Uniform (0, 72) | 168 | 0.238279062 |
| Ebola | Uniform (3, 5) | Uniform (0, 168) | 168 | 0.366941411 |
| Ebola | Uniform (9, 11) | Uniform (0, 168) | 168 | 0.38765292 |
| Ebola | Uniform (15, 17) | Uniform (0, 168) | 168 | 0.414116431 |
| Ebola | Uniform (3, 5) | Uniform (0, 336) | 168 | 0.530061985 |
| Ebola | Uniform (9, 11) | Uniform (0, 336) | 168 | 0.547185583 |
| Ebola | Uniform (15, 17) | Uniform (0, 336) | 168 | 0.568340633 |
| Ebola | Uniform (3, 5) | Uniform (0, 72) | 240 | 0.465677254 |
| Ebola | Uniform (9, 11) | Uniform (0, 72) | 240 | 0.489428097 |
| Ebola | Uniform (15, 17) | Uniform (0, 72) | 240 | 0.512126334 |
| Ebola | Uniform (3, 5) | Uniform (0, 168) | 240 | 0.622570539 |
| Ebola | Uniform (9, 11) | Uniform (0, 168) | 240 | 0.640064404 |
| Ebola | Uniform (15, 17) | Uniform (0, 168) | 240 | 0.660989613 |
| Ebola | Uniform (3, 5) | Uniform (0, 336) | 240 | 0.732647498 |
| Ebola | Uniform (9, 11) | Uniform (0, 336) | 240 | 0.751150595 |
| Ebola | Uniform (15, 17) | Uniform (0, 336) | 240 | 0.764705157 |
| Ebola | Uniform (3, 5) | Uniform (0, 72) | 336 | 0.782056923 |
| Ebola | Uniform (9, 11) | Uniform (0, 72) | 336 | 0.793133451 |
| Ebola | Uniform (15, 17) | Uniform (0, 72) | 336 | 0.807232651 |
| Ebola | Uniform (3, 5) | Uniform (0, 168) | 336 | 0.85698649 |
| Ebola | Uniform (9, 11) | Uniform (0, 168) | 336 | 0.867138891 |
| Ebola | Uniform (15, 17) | Uniform (0, 168) | 336 | 0.877351415 |
| Ebola | Uniform (3, 5) | Uniform (0, 336) | 336 | 0.903144035 |
| Ebola | Uniform (9, 11) | Uniform (0, 336) | 336 | 0.912199334 |
| Ebola | Uniform (15, 17) | Uniform (0, 336) | 336 | 0.917847827 |
| SARS | Uniform (3, 5) | Uniform (0, 72) | 72 | 0.406207969 |
| SARS | Uniform (9, 11) | Uniform (0, 72) | 72 | 0.451983627 |
| SARS | Uniform (15, 17) | Uniform (0, 72) | 72 | 0.496132829 |
| SARS | Uniform (3, 5) | Uniform (0, 168) | 72 | 0.560891103 |
| SARS | Uniform (9, 11) | Uniform (0, 168) | 72 | 0.601420792 |
| SARS | Uniform (15, 17) | Uniform (0, 168) | 72 | 0.638878157 |
| SARS | Uniform (3, 5) | Uniform (0, 336) | 72 | 0.582393083 |
| SARS | Uniform (9, 11) | Uniform (0, 336) | 72 | 0.621546585 |
| SARS | Uniform (15, 17) | Uniform (0, 336) | 72 | 0.653829464 |
| SARS | Uniform (3, 5) | Uniform (0, 72) | 120 | 0.733378854 |
| SARS | Uniform (9, 11) | Uniform (0, 72) | 120 | 0.765099754 |
| SARS | Uniform (15, 17) | Uniform (0, 72) | 120 | 0.796442335 |
| SARS | Uniform (3, 5) | Uniform (0, 168) | 120 | 0.829219814 |
| SARS | Uniform (9, 11) | Uniform (0, 168) | 120 | 0.850658356 |
| SARS | Uniform (15, 17) | Uniform (0, 168) | 120 | 0.869697707 |
| SARS | Uniform (3, 5) | Uniform (0, 336) | 120 | 0.832521431 |
| SARS | Uniform (9, 11) | Uniform (0, 336) | 120 | 0.85715063 |
| SARS | Uniform (15, 17) | Uniform (0, 336) | 120 | 0.875696341 |
| SARS | Uniform (3, 5) | Uniform (0, 72) | 168 | 0.924589337 |
| SARS | Uniform (9, 11) | Uniform (0, 72) | 168 | 0.939448712 |
| SARS | Uniform (15, 17) | Uniform (0, 72) | 168 | 0.949728475 |
| SARS | Uniform (3, 5) | Uniform (0, 168) | 168 | 0.954471475 |
| SARS | Uniform (9, 11) | Uniform (0, 168) | 168 | 0.961202716 |
| SARS | Uniform (15, 17) | Uniform (0, 168) | 168 | 0.96987112 |
| SARS | Uniform (3, 5) | Uniform (0, 336) | 168 | 0.954467704 |
| SARS | Uniform (9, 11) | Uniform (0, 336) | 168 | 0.964453624 |
| SARS | Uniform (15, 17) | Uniform (0, 336) | 168 | 0.970835379 |
| SARS | Uniform (3, 5) | Uniform (0, 72) | 240 | 0.996339263 |
| SARS | Uniform (9, 11) | Uniform (0, 72) | 240 | 0.996963712 |
| SARS | Uniform (15, 17) | Uniform (0, 72) | 240 | 0.998045295 |
| SARS | Uniform (3, 5) | Uniform (0, 168) | 240 | 0.997705114 |
| SARS | Uniform (9, 11) | Uniform (0, 168) | 240 | 0.998259157 |
| SARS | Uniform (15, 17) | Uniform (0, 168) | 240 | 0.998685583 |
| SARS | Uniform (3, 5) | Uniform (0, 336) | 240 | 0.998179051 |
| SARS | Uniform (9, 11) | Uniform (0, 336) | 240 | 0.998216264 |
| SARS | Uniform (15, 17) | Uniform (0, 336) | 240 | 0.998781016 |
| SARS | Uniform (3, 5) | Uniform (0, 72) | 336 | 0.999989504 |
| SARS | Uniform (9, 11) | Uniform (0, 72) | 336 | 1 |
| SARS | Uniform (15, 17) | Uniform (0, 72) | 336 | 1 |
| SARS | Uniform (3, 5) | Uniform (0, 168) | 336 | 1 |
| SARS | Uniform (9, 11) | Uniform (0, 168) | 336 | 1 |
| SARS | Uniform (15, 17) | Uniform (0, 168) | 336 | 0.999985692 |
| SARS | Uniform (3, 5) | Uniform (0, 336) | 336 | 1 |
| SARS | Uniform (9, 11) | Uniform (0, 336) | 336 | 1 |
| SARS | Uniform (15, 17) | Uniform (0, 336) | 336 | 1 |
| Influenza | Uniform (3, 5) | Uniform (0, 72) | 72 | 0.995269971 |
| Influenza | Uniform (9, 11) | Uniform (0, 72) | 72 | 0.997564952 |
| Influenza | Uniform (15, 17) | Uniform (0, 72) | 72 | 0.998650074 |
| Influenza | Uniform (3, 5) | Uniform (0, 168) | 72 | 0.995850015 |
| Influenza | Uniform (9, 11) | Uniform (0, 168) | 72 | 0.997292241 |
| Influenza | Uniform (15, 17) | Uniform (0, 168) | 72 | 0.998468682 |
| Influenza | Uniform (3, 5) | Uniform (0, 336) | 72 | 0.995566939 |
| Influenza | Uniform (9, 11) | Uniform (0, 336) | 72 | 0.997081428 |
| Influenza | Uniform (15, 17) | Uniform (0, 336) | 72 | 0.998313157 |
| Influenza | Uniform (3, 5) | Uniform (0, 72) | 120 | 0.999978951 |
| Influenza | Uniform (9, 11) | Uniform (0, 72) | 120 | 1 |
| Influenza | Uniform (15, 17) | Uniform (0, 72) | 120 | 1 |
| Influenza | Uniform (3, 5) | Uniform (0, 168) | 120 | 1 |
| Influenza | Uniform (9, 11) | Uniform (0, 168) | 120 | 0.999951784 |
| Influenza | Uniform (15, 17) | Uniform (0, 168) | 120 | 1 |
| Influenza | Uniform (3, 5) | Uniform (0, 336) | 120 | 1 |
| Influenza | Uniform (9, 11) | Uniform (0, 336) | 120 | 0.999901681 |
| Influenza | Uniform (15, 17) | Uniform (0, 336) | 120 | 1 |
| Influenza | Uniform (3, 5) | Uniform (0, 72) | 168 | 1 |
| Influenza | Uniform (9, 11) | Uniform (0, 72) | 168 | 1 |
| Influenza | Uniform (15, 17) | Uniform (0, 72) | 168 | 1 |
| Influenza | Uniform (3, 5) | Uniform (0, 168) | 168 | 1 |
| Influenza | Uniform (9, 11) | Uniform (0, 168) | 168 | 1 |
| Influenza | Uniform (15, 17) | Uniform (0, 168) | 168 | 1 |
| Influenza | Uniform (3, 5) | Uniform (0, 336) | 168 | 1 |
| Influenza | Uniform (9, 11) | Uniform (0, 336) | 168 | 1 |
| Influenza | Uniform (15, 17) | Uniform (0, 336) | 168 | 1 |
| Influenza | Uniform (3, 5) | Uniform (0, 72) | 240 | 1 |
| Influenza | Uniform (9, 11) | Uniform (0, 72) | 240 | 1 |
| Influenza | Uniform (15, 17) | Uniform (0, 72) | 240 | 1 |
| Influenza | Uniform (3, 5) | Uniform (0, 168) | 240 | 1 |
| Influenza | Uniform (9, 11) | Uniform (0, 168) | 240 | 1 |
| Influenza | Uniform (15, 17) | Uniform (0, 168) | 240 | 1 |
| Influenza | Uniform (3, 5) | Uniform (0, 336) | 240 | 1 |
| Influenza | Uniform (9, 11) | Uniform (0, 336) | 240 | 1 |
| Influenza | Uniform (15, 17) | Uniform (0, 336) | 240 | 1 |
| Influenza | Uniform (3, 5) | Uniform (0, 72) | 336 | 1 |
| Influenza | Uniform (9, 11) | Uniform (0, 72) | 336 | 1 |
| Influenza | Uniform (15, 17) | Uniform (0, 72) | 336 | 1 |
| Influenza | Uniform (3, 5) | Uniform (0, 168) | 336 | 1 |
| Influenza | Uniform (9, 11) | Uniform (0, 168) | 336 | 1 |
| Influenza | Uniform (15, 17) | Uniform (0, 168) | 336 | 1 |
| Influenza | Uniform (3, 5) | Uniform (0, 336) | 336 | 1 |
| Influenza | Uniform (9, 11) | Uniform (0, 336) | 336 | 1 |
| Influenza | Uniform (15, 17) | Uniform (0, 336) | 336 | 1 |
| COVID-19 | Uniform (3, 5) | Uniform (0, 72) | 72 | 0.416056894 |
| COVID-19 | Uniform (9, 11) | Uniform (0, 72) | 72 | 0.459551256 |
| COVID-19 | Uniform (15, 17) | Uniform (0, 72) | 72 | 0.500688592 |
| COVID-19 | Uniform (3, 5) | Uniform (0, 168) | 72 | 0.523042637 |
| COVID-19 | Uniform (9, 11) | Uniform (0, 168) | 72 | 0.564279419 |
| COVID-19 | Uniform (15, 17) | Uniform (0, 168) | 72 | 0.598082762 |
| COVID-19 | Uniform (3, 5) | Uniform (0, 336) | 72 | 0.543501801 |
| COVID-19 | Uniform (9, 11) | Uniform (0, 336) | 72 | 0.583228533 |
| COVID-19 | Uniform (15, 17) | Uniform (0, 336) | 72 | 0.619733953 |
| COVID-19 | Uniform (3, 5) | Uniform (0, 72) | 120 | 0.706608256 |
| COVID-19 | Uniform (9, 11) | Uniform (0, 72) | 120 | 0.73368012 |
| COVID-19 | Uniform (15, 17) | Uniform (0, 72) | 120 | 0.757190601 |
| COVID-19 | Uniform (3, 5) | Uniform (0, 168) | 120 | 0.770193737 |
| COVID-19 | Uniform (9, 11) | Uniform (0, 168) | 120 | 0.78877531 |
| COVID-19 | Uniform (15, 17) | Uniform (0, 168) | 120 | 0.810140719 |
| COVID-19 | Uniform (3, 5) | Uniform (0, 336) | 120 | 0.779018476 |
| COVID-19 | Uniform (9, 11) | Uniform (0, 336) | 120 | 0.796699836 |
| COVID-19 | Uniform (15, 17) | Uniform (0, 336) | 120 | 0.817129102 |
| COVID-19 | Uniform (3, 5) | Uniform (0, 72) | 168 | 0.865829451 |
| COVID-19 | Uniform (9, 11) | Uniform (0, 72) | 168 | 0.87869567 |
| COVID-19 | Uniform (15, 17) | Uniform (0, 72) | 168 | 0.891054904 |
| COVID-19 | Uniform (3, 5) | Uniform (0, 168) | 168 | 0.894395917 |
| COVID-19 | Uniform (9, 11) | Uniform (0, 168) | 168 | 0.905888281 |
| COVID-19 | Uniform (15, 17) | Uniform (0, 168) | 168 | 0.915047219 |
| COVID-19 | Uniform (3, 5) | Uniform (0, 336) | 168 | 0.90086556 |
| COVID-19 | Uniform (9, 11) | Uniform (0, 336) | 168 | 0.912036804 |
| COVID-19 | Uniform (15, 17) | Uniform (0, 336) | 168 | 0.916215403 |
| COVID-19 | Uniform (3, 5) | Uniform (0, 72) | 240 | 0.959891894 |
| COVID-19 | Uniform (9, 11) | Uniform (0, 72) | 240 | 0.963770202 |
| COVID-19 | Uniform (15, 17) | Uniform (0, 72) | 240 | 0.965725392 |
| COVID-19 | Uniform (3, 5) | Uniform (0, 168) | 240 | 0.968058692 |
| COVID-19 | Uniform (9, 11) | Uniform (0, 168) | 240 | 0.970998624 |
| COVID-19 | Uniform (15, 17) | Uniform (0, 168) | 240 | 0.975199989 |
| COVID-19 | Uniform (3, 5) | Uniform (0, 336) | 240 | 0.972098299 |
| COVID-19 | Uniform (9, 11) | Uniform (0, 336) | 240 | 0.971414887 |
| COVID-19 | Uniform (15, 17) | Uniform (0, 336) | 240 | 0.975229704 |
| COVID-19 | Uniform (3, 5) | Uniform (0, 72) | 336 | 0.9916526 |
| COVID-19 | Uniform (9, 11) | Uniform (0, 72) | 336 | 0.991684312 |
| COVID-19 | Uniform (15, 17) | Uniform (0, 72) | 336 | 0.99313047 |
| COVID-19 | Uniform (3, 5) | Uniform (0, 168) | 336 | 0.992592385 |
| COVID-19 | Uniform (9, 11) | Uniform (0, 168) | 336 | 0.994013238 |
| COVID-19 | Uniform (15, 17) | Uniform (0, 168) | 336 | 0.994013991 |
| COVID-19 | Uniform (3, 5) | Uniform (0, 336) | 336 | 0.993342076 |
| COVID-19 | Uniform (9, 11) | Uniform (0, 336) | 336 | 0.994046117 |
| COVID-19 | Uniform (15, 17) | Uniform (0, 336) | 336 | 0.99458814 |
